# The Impact of the COVID-19 Outbreak on the Medical Treatment of Chinese Children with Chronic Kidney Disease (CKD) : A Multicenter Cross-section Study in the Context of a Public Health Emergency of International Concern

**DOI:** 10.1101/2020.02.28.20029199

**Authors:** Gaofu Zhang, Haiping Yang, Aihua Zhang, Qian Shen, Li Wang, Zhijuan Li, Yuhong Li, Lijun Zhao, Yue Du, Liangzhong Sun, Bo Zhao, Hongtao Zhu, Haidong Fu, Xiaoyan Li, Xiaojie Gao, Sheng Hao, Juanjuan Ding, Zongwen Chen, Zhiquan Xu, Xiaorong Liu, Daoqi Wu, Mingsi Gao, Mo Wang, Qiu Li

**Author notes:** **Correspondence to:** Dr Qiu Li;, Dr Mo Wang.

## Abstract

**Objective:** To investigate the impact of the COVID-19 outbreak on the medical advice seeking of Chinese children with chronic kidney disease (CKD).

**Materials and Methods:** An anonymous online questionnaire survey was conducted in 17 pediatric nephropathy diagnosis and treatment centers in China. The questions collected basic information on the patients and their parents and data on changes in the approach to medical treatment and their needs in the context of the outbreak etc. This is a Multicenter Cross-section Study.

**Results:** A total of 735 valid questionnaires were collected. 555 patients (75.5%) and their parents said that the outbreak had a significant influence on their medical treatment: 264 patients (47.6%) said that it would be delayed by 2 to 4 weeks and 199 patients (35.9%) by 4 to 8 weeks. 510 patients (84.16%) hoped to get in touch with specialists through online consultation, and 528 patients (84.5%) hoped that online consultation could be implemented and that medication could be delivered to them.. A total of 458 patients (62.3%) said that their greatest concern was that the CKD would be aggravated or that they would experience a relapse; only 203 patients were infected by 2019-nCoV. A total of 313 patients (42.5%) experienced anxiety and thus required the intervention of psychologists.

**Conclusion:** The COVID-19 outbreak has affected the medical treatment of children with CKD. Online consultation, medication delivery and psychological counselling are the greatest needs reported by patients and their families and could especially provide solutions for the management of low income children with CKD in remote rural areas in the context of the COVID-19 epidemic.

**Strengths and limitations of this study:** The study is a Multicenter Cross-section Study in the Context of a Public Health Emergency of International Concern.

The study can well explore the impact of COVID-19 outbreak on the medical treatment of CKD children in China and the needs of current patients.

The study explored ways to meet the medical needs of CKD children in the context of a public health emergency of international concern, which provides a method support for all countries in the world experiencing COVID-19 outbreak.

The study is an exploration of the coping strategies for CKD management in China.

The limitations of this study is that it is quite simple and descriptive, and many studies need to be further carried out.

## Introduction

In December 2019, multiple cases of pneumonia with unknown causes were identified in Wuhan, Hubei Province; The patients were soon confirmed to have been infected by a new coronavirus. On January 12, 2020, the World Health Organization (WHO) officially named it the “2019 novel coronavirus (2019-nCoV)”. On January 30, 2020, the WHO declared the outbreak of 2019-nCoV in China to be a “public health emergency of international concern”. On February 11, 2020, the WHO announced the official name of the disease caused by 2019-nCoV: COVID-19 (Corona Virus Disease 2019, COVID-19). In addition, the International Committee on Taxonomy of Viruses (ICTV) announced the official classification of 2019-nCoV as severe acute respiratory syndrome coronavirus 2 (SARS-CoV-2)[1-4]. By 0:00 on February 17, 2020, COVID-19 had spread to all provinces and cities in China and more than 20 foreign countries, with 68,595 confirmed cases, 8,228 suspected cases, 9,755 cured cases and 1,667 deaths in China and cases among children continuing to rise[5]. Epidemiological investigations have shown that the population is generally susceptible to SARS-CoV-2, and the elderly and patients with underlying diseases become more seriously ill after being infected, with a poor prognosis and high mortality[6]. Because of this public health emergency of international concern, the Chinese government has taken a series of measures to control the epidemic, including sealing off cities, controlling roads and vehicles, and enforcing home quarantine[7-8]. Children with chronic kidney disease (CKD) are a high-risk group highly susceptible to SARS-CoV-2 due to underlying disease and poor immunity[9-10].

CKD in children refers to chronic structural and/or functional abnormalities of the kidney due to various causes lasting for more than 3 months and can eventually develop into end-stage renal disease (ESRD)[11]. CKD has a high incidence worldwide and has become one of the major public health problems affecting children. At present, the treatment of children with stage II CKD mainly involves the treatment of the primary disease. After Stage II (when the glomerular filtration rate falls below 90 ml/min/1.73 m^2^), long-term comprehensive management is carried out, and efforts are made to detect the causes, remove or reduce the damage factors, delay the progressive damage of renal function and actively treat and reduce complications before the disease develops to Stage V[11-12]. The treatment of children with stage V CKD mainly involves renal replacement therapies (including kidney transplantation, maintenance blood purification and peritoneal dialysis) and symptomatic treatment of complications[11-12]. Therefore, all children with CKD need to travel between home and the hospital at different frequencies and have a high requirement for the continuity of medical treatment.

In the context of the public health emergency of international concern of the COVID-19 outbreak, the management and medical treatment of children with CKD and the mental status of their parents will certainly be affected. To prevent and control SARS-CoV-2 infection, continue the long-term standard management of children with CKD, better serve children with CKD and avoid the spread of the epidemic, we conducted an anonymous online questionnaire survey in collaboration with the 12 largest pediatric nephropathy diagnosis and treatment centers in China. Through the survey, we sought to understand the effect of the SARS-CoV-2 outbreak on the medical treatment of various kinds of Chinese children with CKD, changes in approaches to their medical treatment, their greatest needs at present and changes in the mental status of their parents to explore problems exposed in the management of Chinese children with CKD in the context of a public health emergency of international concern and identify coping strategies.

## Materials and Methods

### Study design and participant recruitment

A cross-sectional study based on an online questionnaire survey was conducted between 0:00, February 10^th^, 2020, and 0:00, February 14^th^, 2020, in pediatric nephropathy diagnosis and treatment centers in 17 provinces and municipalities in China. The 17 centers are located in such regions as northeastern China, eastern China, southern China, central China, western China and northwestern China, and each of them is the largest pediatric nephropathy diagnosis and treatment center in the province or municipality. All CKD diagnoses met the KIDGO diagnostic criteria[11]. All the patients recruited, screened, and included were inpatients registered at various diagnostic and treatment centers. Questionnaires were sent to the parents of a total of 754 patients with definitive diagnoses via QR codes on WeChat, and 735 valid questionnaires were collected.

### Questionnaire and data collection

This online questionnaire had three parts: the first part collected the basic information of the sick children and their parents (age, gender, diagnosis (CKD stage), address, family economic status, disease duration, attention paid by the parents to the epidemic); the second part assessed the impact of the epidemic on the medical treatment of the patients and their needs for medical treatment (the hospital that they visit, transportation mode, the effect of the epidemic on regular medical treatment, delays in medical treatment, the self-perceived effect of the epidemic on the condition of the sick child, measures that the parents hoped could be taken if they could not see a doctor, help that the parents hoped to get during the epidemic); the third part evaluated the impact of the epidemic on the mental status and anxiety of the parents (mentality of the parents and the attention paid to the COVID-19 outbreak, self-perceived probability of the child being infected by SARS-CoV-2, concern of the parents about the epidemic, anxiety of the parents, the parents’ needs for psychological counseling and whether the parents had received psychological counseling.)

### Statistical analysis

This is a descriptive study.

## Results

### Basic patient information

The questionnaires were given to a total of 754 patients, and 735 valid questionnaires were collected, for a recovery rate of 97.5%. The parents of 735 sick children participated in the survey, of whom 447 were boys (60.8%) and 288 were girls (39.2%). The age distribution was as follows: 63 patients were younger than 3 years old (8.6%), 160 were between 3 and 6 years old (21.8%), 277 were between 6 and 11 years old (37.7%), and 235 were older than 11 years (31.97%) (Fig. 1. B). The distribution of the time since receiving a definitive diagnosis of CKD was as follows: 83 patients had been diagnosed for fewer than 3 months (11.3%), 64 for 3 to 6 months (8.71%), 89 for 6 to 12 months (12.1%), 310 for 1 to 3 years (42.18%) and 189 for more than 3 years (25.7%). Among them, 659 patients had stage I-II CKD (89.6%), 51 had stage III-IV CKD (6.9%), and 25 had stage V CKD and were currently receiving regular blood purification or peritoneal dialysis (3.4%).

**Fig. 1.**
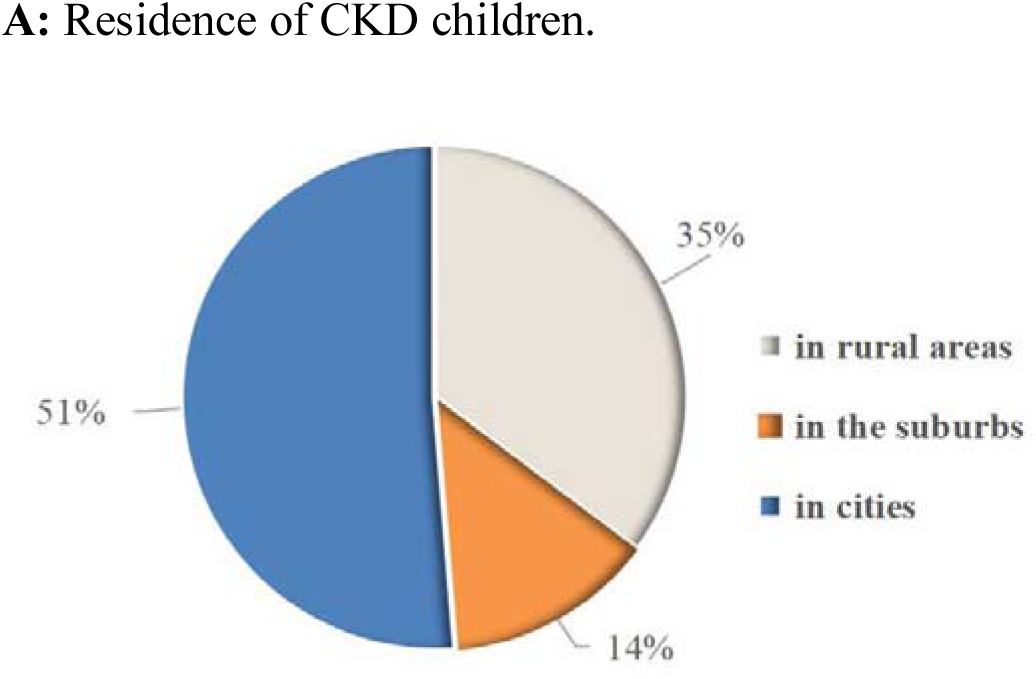
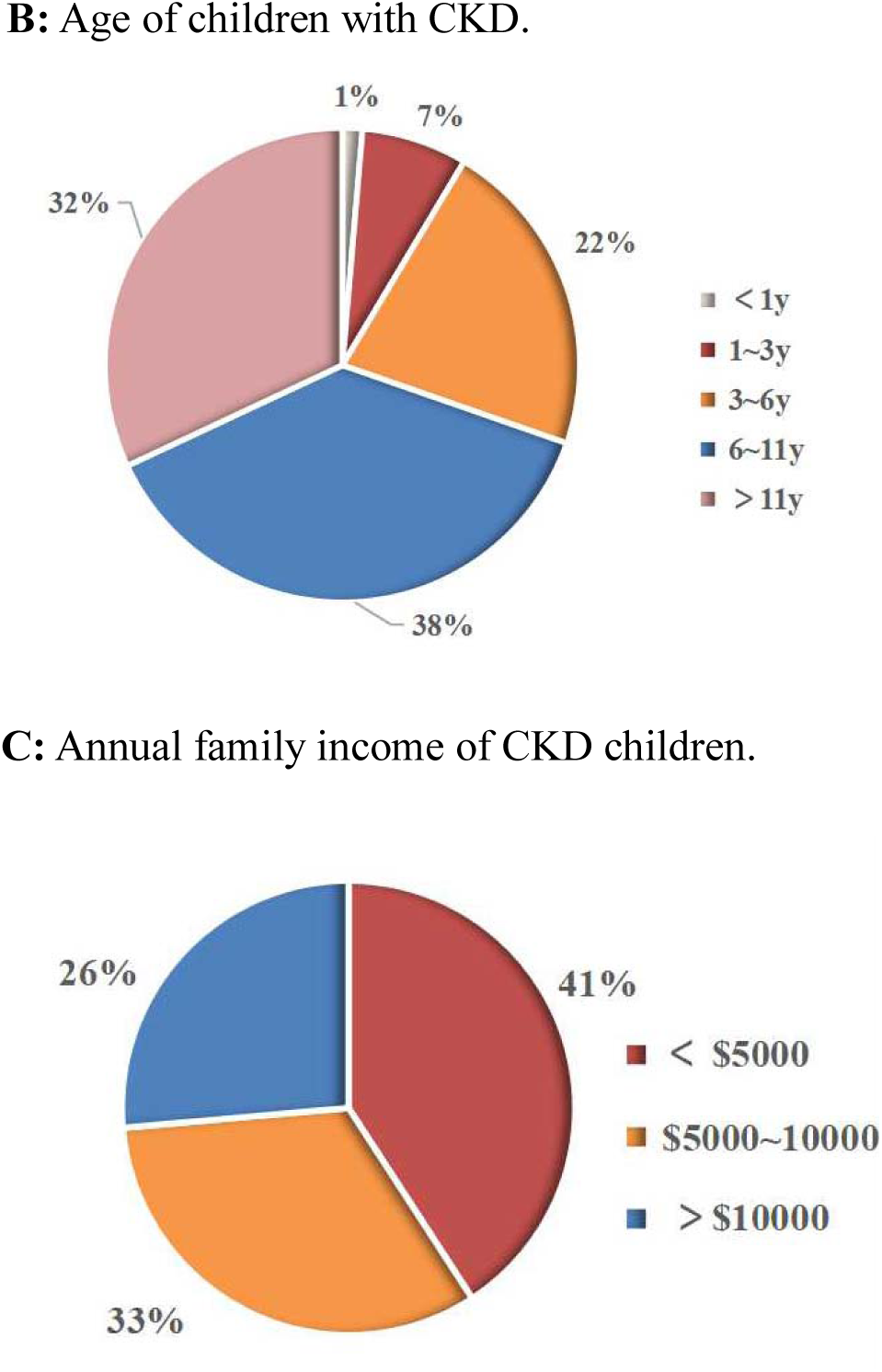
Basic patient information of Chinese children with chronic kidney disease (CKD).

A total of 259 patients (35.2%) lived in rural areas, 99 (13.5) lived in the suburbs and 377 (51.3%) lived in cities. Further investigations revealed that most of those living in cities lived in county-level cities, 235 of whom (62.3%) needed to travel for two hours or more to see a doctor, a lengthy trip (Fig. 1. A).

In terms of parental education level, 345 (46.9%) had a junior high school or lower education, 276 (37.6%) had a senior high school or vocational college education, and 114 (15.5%) had an undergraduate or postgraduate education. In terms of family income, 300 (40.8%) had an annual income of less than $5000, 241 (32.8%) had an annual income of $5000-10000, and 194 (26.3%) had an annual income of more than $10000 (Fig. 1. C)..

### Impact of the epidemic on the medical treatment of the patients and the needs of the parents

Among the parents of the 735 sick children, the parents of 555 patients (75.5%) said that the COVID-19 outbreak had a significant impact on the regular medical treatment of their children: 264 (47.6%) said that it would be delayed by 2 to 4 weeks, 199 (35.9%) said that it would be delayed by 4 to 8 weeks, and 92 (16.6%) even said that it would be delayed by more than 8 weeks. A total of 180 parents (24.5%) said that the impact of the COVID-19 outbreak on the regular medical treatment of their sick child was not significant. Due to reasons such as road control and traffic conditions, 429 parents had difficulties visiting hospitals. Three hundred fifty-two parents (82.1%) said that failing to see the doctor would have an impact on the condition and treatment of their child, and 193 (42.1%) thought that it would have a significant impact(Fig. 2).

**Fig. 2.**
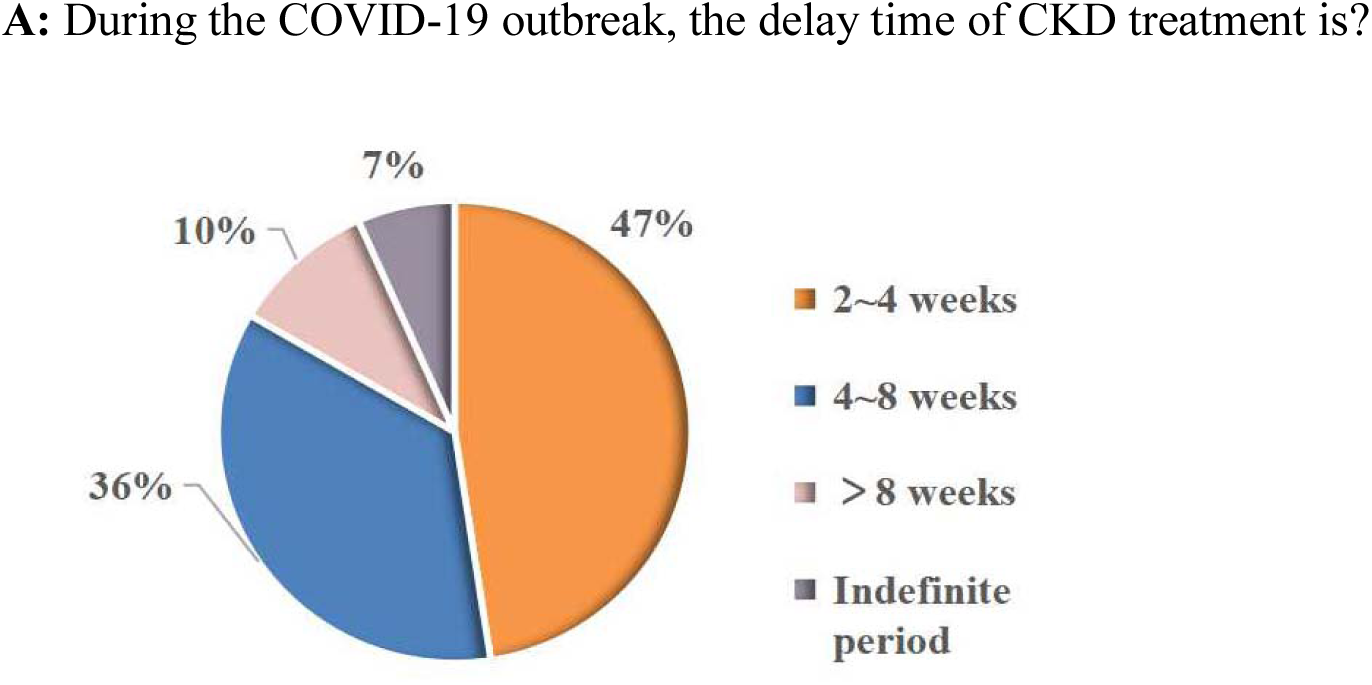
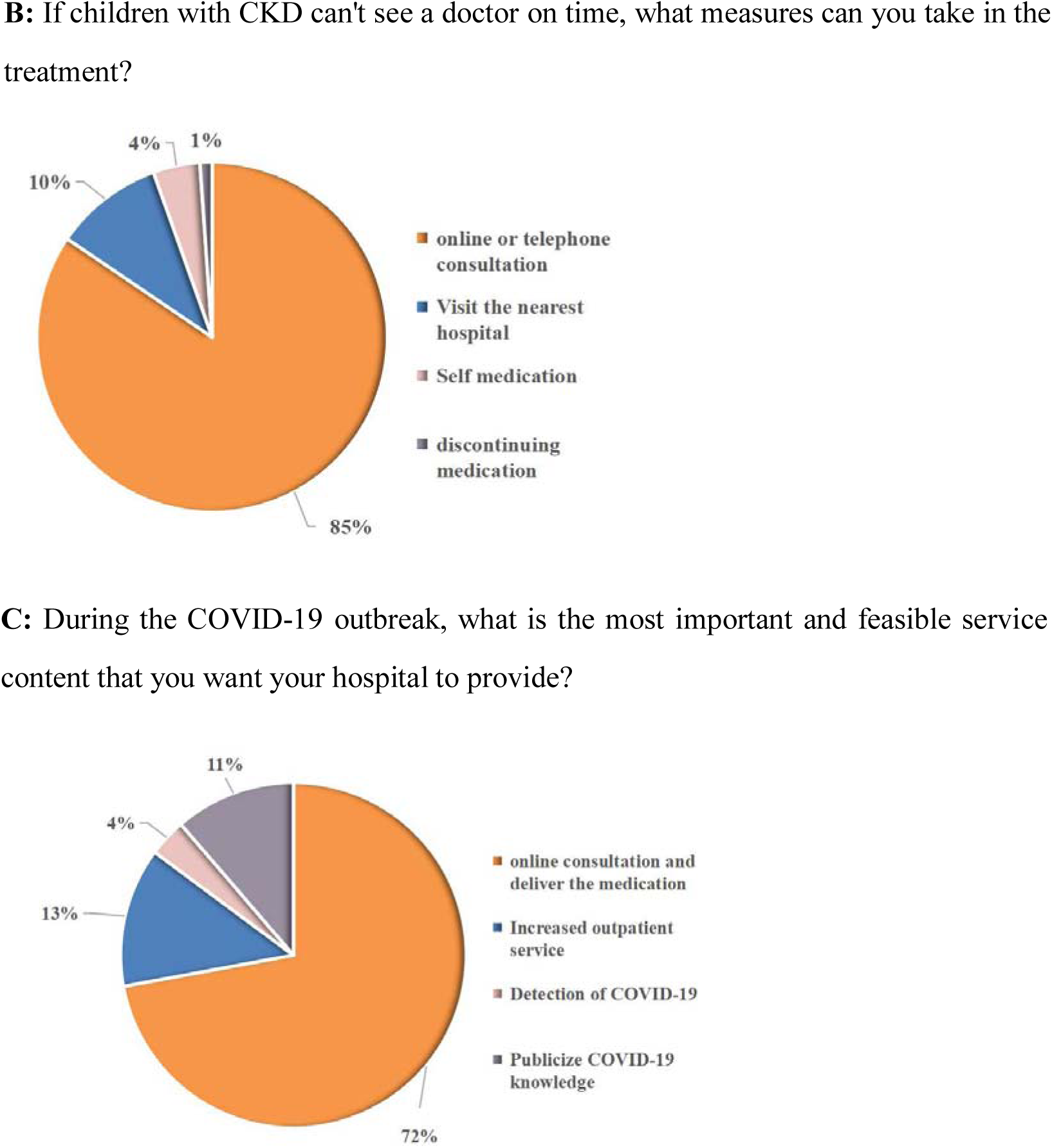
Impact of the epidemic on the medical treatment of the patients and the needs of the parents of Chinese children with chronic kidney disease (CKD).

Because of the COVID-19 epidemic, 84.5% of the parents hoped to get in touch with their nephrologists through online or telephone consultation to discuss the kidney disease and the COVID-19 epidemic; 72.1% of the parents hoped that hospitals could deliver the medication, i.e., with an electronic prescription from the doctor. Eighty-one percent of those hoping the medicines could be delivered were from rural areas or were patients from other provinces(Fig. 2).

### Impact of the epidemic on the mental status and anxiety of the parents of sick children

Among the parents of the 735 sick children, 479 (65.2%) paid attention to the epidemic, spending 30 minutes to 1 hour each day following news of the epidemic and learning relevant medical knowledge. One hundred seventy-eight parents (24.2%)paid much attention to the news of the epidemic and spent more than 1 hour each day learning relevant knowledge on the prevention and treatment of SARS-CoV-2(Fig. 3).

**Fig. 3.**
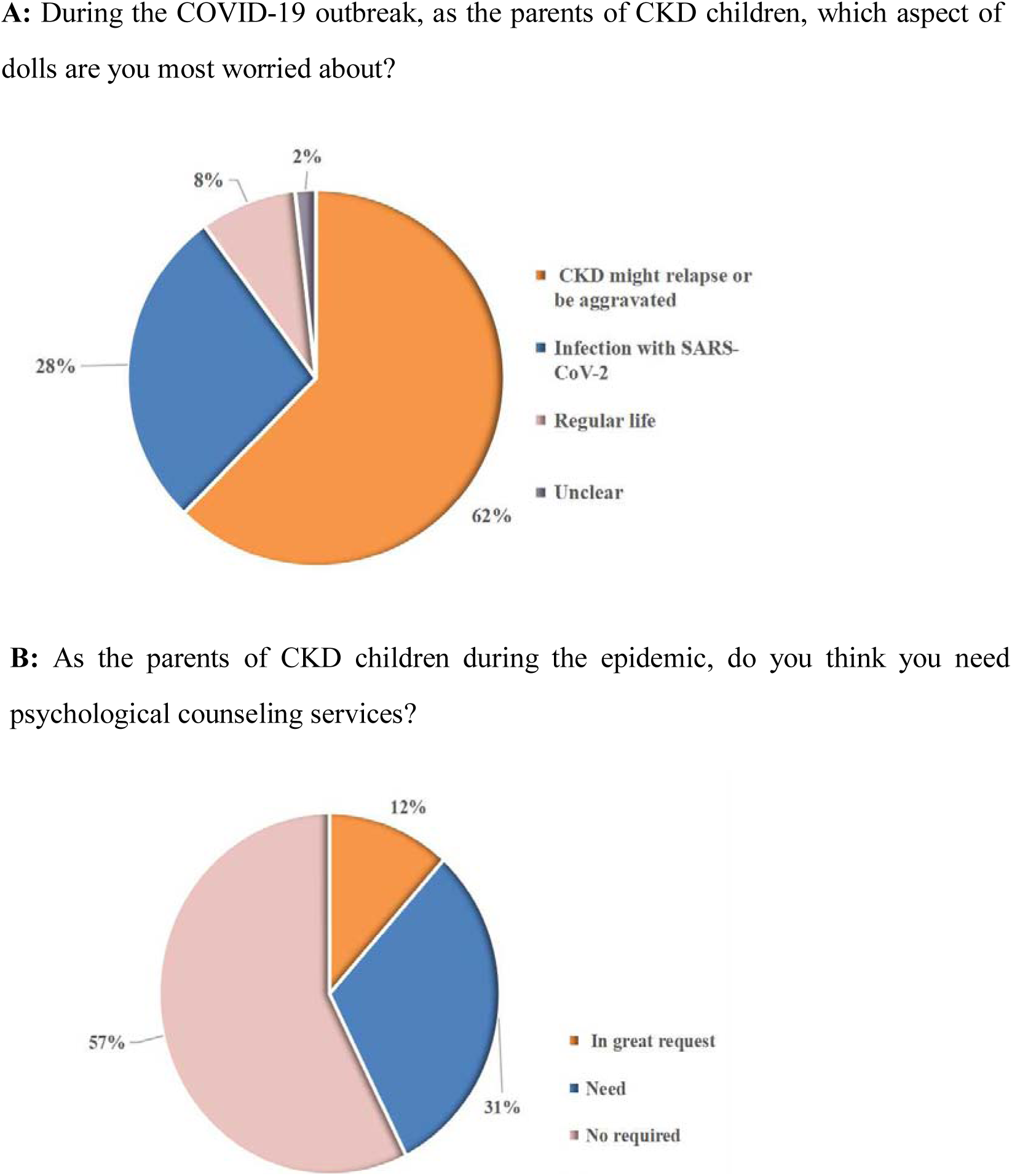
Impact of the epidemic on the mental status and anxiety of the parents of Chinese children with chronic kidney disease (CKD).

Due to the COVID-19 epidemic, 594 parents (80.8%) were concerned about the condition of their sick children, 532 of whom (72.4%) said that children with kidney diseases were more susceptible to SARS-CoV-2 because of their low immunity. A total of 496 parents (67.5%) were more anxious than the general population, worrying about aggravation of the original kidney disease or infection with SARS-CoV-2. In terms of anxiety, 458 parents (62.3%) were worried that the CKD might relapse or be aggravated during the epidemic since they could not regularly see the doctor; 203 parents (27.6%) were concerned about being infected by SARS-CoV-2 during the epidemic. A total of 313 parents (42.5%) hoped to receive psychological counselling, and 409 parents (75.5%) said that there were no channels for them to receive psychological counselling(Fig. 3).

## Discussion

On January 21, 2020, the National Health Commission of China decided to include COVID-19 pneumonia as a national “class B” infectious disease and manage it as a “class A” infectious disease[5,7]. In the context of this public health emergency of international concern, many problems in the management of Chinese children with CKD have been exposed, and we sought to explore solutions to these issues.

In our survey, we found that 48.7% of the patients lived in remote rural areas or suburbs, making it inconvenient for them to see a doctor. Although 51.3% lived in cities, most of them lived in county-level cities and needed to travel to provincial capitals or municipalities directly under the central government for medical treatment. Despite the inconvenience of medical treatment, only 14.2% of the patients would choose to receive medical treatment, reexaminations and follow-up visits in district or county-level hospitals, and the other 85.8% preferred to have reexaminations and follow-up visits in grade-A class-3 hospitals and large specialized hospitals. According to the opinion of the parents, due to the lack of pediatric nephropathy knowledge, relevant examinations for kidney diseases and medication, district and county-level hospitals could not meet their needs for medical treatment, revealing a problem related to a lack of diagnosis and treatment standards for CKD in basic pediatric departments. As a result, some common kidney diseases cannot be addressed in local hospitals. A complete three-level system for the specialized diagnosis and treatment of children with CKD should be established.

In our survey, we found that although 81.1% of the parents were paying attention to the COVID-19 epidemic and the situation of the CKD itself, only 4.32% planned to adjust the medication dosage based on their previous medical knowledge and experience. In fact, 1.12% of the parents said they would directly stop the medicines or end the treatment, and 84.5% said that they would rely more on the guidance of doctors, which indicates the low participation of children with CKD and their families. In the absence or interruption of the guidance of medical teams, disruptions may occur in the management of the disease. Similarly, we found in our survey that in the face of this serious epidemic, 76.7% of the parents of children with CKD showed a lack of prevention and infection control knowledge, as well as knowledge of channels for nutrition guidance and emergency coping and appeal making; 72.1% of the parents said that they lacked access to medical treatment and medication, suggesting that in major public health emergencies, the Chinese medical service system should give full play to the unique advantages of online hospitals and online diagnosis and treatment, encourage online follow-up consultation for some common diseases and chronic diseases, and reduce the risk of cross-infection among patients in offline treatment. Doctors can carry out online consultations and provide electronic prescriptions, and the patients can then pay the fee for the consultation and medication online. Payment can then be verified by doctors and pharmacists online, and after successful payment, the orders can be taken via electronic prescriptions at pharmacies. In the future, online hospitals should also improve their emergency prescription-taking measures. For example, during major epidemics, cooperation and contact with logistics enterprises can be strengthened to deliver medication to the homes of the patients, indicating that we need a stable online “internet + CKD health management” platform and online hospital systems to cope with public health emergencies of international concern.

We found in our survey that 67.5% of the parents were more anxious than the general population due to worry that the primary kidney disease would be aggravated or that they might be infected with SARS-CoV-2. Among them, 42.5% of the parents expressed that they hoped to receive psychological counselling, but the access of children with CKD and their parents to counselling was blocked, leaving them isolated and helpless and resulting in psychological anxiety, helplessness and even panic. Therefore, in the context of a public health emergency of international concern, the psychological status of children with CKD and their family members should first be determined to identify high-risk groups and avoid such extreme events as suicide and impulsiveness. Therefore, psychological services and interventions for the mental health of children with CKD and their families at early stages are needed. For example, during this COVID-19 outbreak, it is necessary to provide children with CKD and their family members with psychological support and psychological counselling, thus preventing and reducing frustration caused by the epidemic and preventing extreme events caused by mental stress. To this end, a hotline for psychological guidance can be set up, and guidelines for this hotline should be formulated to effectively alleviate the tension and panic of the public.

## Conclusion

During this COVID-19 outbreak, many problems in the management of Chinese children with CKD have been exposed, indicating the need to establish a regional contingency plan for CKD management in the context of major public health emergencies. Considering the characteristics of CKD as a chronic disease, a three-level diagnosis and treatment system for children with chronic diseases such as CKD should be established to give full play to graded diagnosis and bidirectional transfer. To gradually improve the care system for children with chronic diseases such as CKD, it is necessary to improve the service function and operation mechanism of “internet+ CKD health management”, including health assessment of children with chronic diseases, health guidance, health education, medical guidance, and psychological counselling, through service applications such as follow-up consultation, diagnosis and treatment appointments, online follow-up visits, adjustment of disease management plans, medication delivery and health management.

## Data Availability

All data is available during the study appear in the submitted article.

## Acknowledgments

Thanks to all the parents of CKD children who participated in the questionnaire.

## Author Contributions

Gaofu Zhang and Yang Haiping are co-first authors. Mo Wang and Qiu Li are corresponding author. Study Design: Mo Wang, Qiu Li; Data Collection: Gaofu Zhang, Haiping Yang, Aihua Zhang, Qian Shen, Li Wang, Zhijuan Li, Yuhong Li, Lijun Zhao, Yue Du, Liangzhong Sun, Bo Zhao, Hongtao Zhu, Haidong Fu, Xiaoyan Li, Xiaojie Gao, Sheng Hao, Juanjuan Ding, Zongwen Chen, Zhiquan Xu, Xiaorong Liu;Statistical Analysis: Gaofu Zhang, Haiping Yang, Data Interpretation: Mo Wang, Qiu Li;Literature Search: Daoqi Wu, Mingsi Gao. All authors contributed to the development of the manuscript, and read and endorsed its final version.

## Author Approval

All authors have seen and approved the manuscript.

## Data sharing

No additional data are available.

## Ethics approval

The study protocol is approved by Ethics Committee of Children’s Hospital of Chongqing Medical University.

